# Genetic co-occurrence networks identify polymorphisms within ontologies highly associated with preeclampsia

**DOI:** 10.1101/2024.07.09.24310164

**Authors:** Andreea Obersterescu, Brian J. Cox

## Abstract

Polygenic diseases require the co-occurrence of multiple risk variants to initiate a pathology. An example is preeclampsia, a hypertensive disease of pregnancy with no known cure or therapy other than the often-preterm delivery of the neonate and placenta. Preeclampsia is challenging to predict due to symptomatic and outcome heterogeneity. Transcriptomic and genetic analysis suggests that this is a multi-syndromic and multigenic disease. Previous research applications of traditional GWAS methods to preeclampsia identified only a few alleles with marginal differences between cases and controls. We seek to identify genetic networks related to the pathophysiology of preeclampsia as potential avenues of therapeutic investigation and early genetic testing. We created a novel systems biology-based method that identifies networks of co-occurring SNPs associated with a trait or disease. The co-occurring pairs are assembled into higher-order associations using network graphs. We validated our method using simulation modelling and tested it against maternal genetic data of a previously assessed preeclampsia cohort. The genetic co-occurrence network identified SNPs in or near genes with ontological enrichment for VEGF, immunological and hormonal pathways, among others with known physiological disruption in preeclampsia. Our findings suggests that preeclampsia is caused by relatively common alleles (<5%) that accumulate in unfavorable combinations.

## Introduction

Preeclampsia is a hypertensive disorder of pregnancy that globally causes over 70,000 maternal deaths and 500,000 neonatal deaths annually^1^. Preeclampsia is largely untreatable aside from medically induced delivery to limit fetal and maternal mortality^2^. In high-resource settings, it accounts for $10s of billion in health care costs annually and a large proportion of maternal and neonatal admissions to hospital and intensive care. Preeclampsia is challenging to detect before the onset of overt symptoms of hypertension and kidney dysfunction (proteinuria), and the disease can progress rapidly^3,4^. Most early detection methods, such as sFLT/PGF ratios, have high negative predictive value to help rule out people^5^. Symptomatic onset is variable, occurring between week 24 and term^6^. The preeclampsia pathology is multifactorial, with most cases presenting with maternal systemic inflammation of the vascular system (hypertension) and inflammation of the liver, kidney and heart^7,8^. On the fetal side, there can be fetal and placental growth restriction. Most frequently discussed are various pathologies involving defective angiogenesis and vascular remodelling^6,9^. Others more strongly favour immunological and metabolic origins of the disease^10,11^. There are likely many subtypes of the disease, and our previous work identified major histological and transcriptional subtypes involving placental development, maternal cardiovascular and immune systems^12–14^.

The maternal and fetal genomes appear to potentially contribute to disease risk, with an estimated contribution of over 50% ^15^. Genetic testing is sought to identify early disease markers and targets for therapeutic intervention and prevention. Genome-wide association studies are generally used to find polymorphisms (SNPs) associated with a disease. GWAS studies on preeclampsia find few associated SNPs and do not replicate well between populations^15^. Maternal studies identified SNPs in the INHBB gene in both Australian and Han Chinese cohorts but not in a Norwegian/Finnish cohort^16,17^. Inhibins are thought to play a role in spiral artery remodelling critical to proper placental development, oxygenation and nutrient transfer ^16^. In an Icelandic/UK cohort, SNPs near the FLT1 gene were found to be associated with PE in a GWAS of the placental (fetal) genome^18^. The FLT1 gene encodes the vascular endothelial growth factor receptor that binds vascular endothelial growth factor alpha (VEGFA). In fetal trophoblasts, the soluble sFLT1 isoform is expressed and released into maternal circulation at levels higher on average than preeclampsia^19^. It is thought that circulating sFLT-1 binds VEGF A (VEGFA), preventing activation of the transmembrane receptor and inhibiting angiogenesis. While these may relate to the common vascular pathologies observed, the association of the SNP with the disease is weak, representing a minority of cases, as many healthy pregnancies carry the alleles.

Moving away from univariate testing and single gene associations to disease, we propose that PE is a polygenic disease resulting from an accumulation of co-occurring SNPs within genes belonging to pathways critical to the observed pathophysiology. The polygenic risk score (PRS) is a current method to address polygenic disease^20^. These scores are created by conducting a GWAS analysis and using the statistical test results to rank and select SNPs based on an arbitrary cut-off, leading to models with dozens to 1000s of SNPs. The prediction is made by weighting SNPs by their effect sizes and summing them^20^. PRS methods highlight that having more SNPs associated with a trait will increase the risk of developing that trait. However, it should be noted that not all SNPs included in the score are biologically relevant, and consequently, scores with fewer SNPs can perform better than those with many^20^. Accepting single polymorphisms above the statistical threshold inflates false discovery and diminishes the ability to discover gene set enrichments. Polymorphisms that contribute to disease are likely to appear in multiple biochemical pathways^21,22^. However, PRSs still utilize a univariate signal and do not consider how many pathways or how severely pathways are hit.

We propose that an improvement is to utilize a framework similar to a synthetic lethal screen, where two or more genes are disrupted simultaneously^23,24^. If the two co-occurring mutations create a phenotype, these are scored as a gene-gene interaction^23,24^. These interacting genes are typically enriched in protein-protein interactions and may be in the same pathway^24^. Alternatively, genetic interactions may occur in parallel pathways such that there is a loss of redundancy^24^.

To address the limitation of GWAS and the PRS methods, we created a novel method that first tests for SNP co-occurrence and then tests for the enrichment of the co-occurrence to a pathology or trait. Importantly, individual SNPs are not strictly required to be enriched to the trait. Co-occurring SNPs enriched to a trait are assembled into higher order structures using network graphs revealing a complexity of genetic interaction associated with preeclampsia. These larger networks are then tested using ontology enrichment to identify key pathways with enriched polymorphic variants in preeclampsia.

## Results

### Simulated random genomes show low background co-occurrence of SNPs

We used data modeling to establish a ground truth data set to evaluate if co-occurrence could used identify polymorphic variants associated with disease. The R package *minutest* was used to simulate 6,000 individuals with 4000 alleles through random breeding. Linkage disequilibrium was simulated at values like those observed in the 1000 genomes project^25^. First, we aimed to establish the background false discovery rate of SNP cooccurrence using simulated small genomes where alleles could be spiked to associate with the disease. To establish the background rate, we randomly drew 1000 individuals from the population for cases and controls each (without replacement). We did so ten times (with replacement) to form paired datasets for ten trials. These should have no true differential co-occurrence.

Since the cohorts comprised 4000 alleles, each trial tested 7,998,000 SNP pairs for co-occurrences. An average of just 3 SNP pairs passed the co-occurrence filter (FDR-adjusted p-value ≤ 0.05 from a hypergeometric test for co-occurrence and FDR-adjusted p-value from a chi-squared test for co-occurrence in case vs control) used to create networks. Additionally, after spiking independent SNPs (either with disease or in LD models), paired datasets had no individual SNPs that were associated with disease (p-value ≤ 5 x 10^-8^ from Cochrane-Armitage trend test). These findings established that our methods produced a low background rate of co-occurring SNPs in the simulations.

### Co-occurring SNPs in cases, controls are separable by graph properties

Next, we aimed to identify the true discovery rate, sensitivity and specificity in simulated small genomes. Additionally, we aimed to determine if network graph properties could be used to identify true discoveries. We used the case-control cohort we generated above to simulate a disease state with co-occurrence (Fig. 1A). In the cases, a range of spiked SNPs (5-80) were selected and forced to correlate at different phi values (0.2, 0.3 and 0.4). Critically, when spiking co-occurring SNPs we did not alter the frequency of the SNPs between the cases and controls ensuring that no individual SNPs was enriched. Each combination of spiked alleles and phi values was evaluated in 10 trials of 1000 cases and 1000 controls and we evaluated their means and distributions.

**Figure 1.**
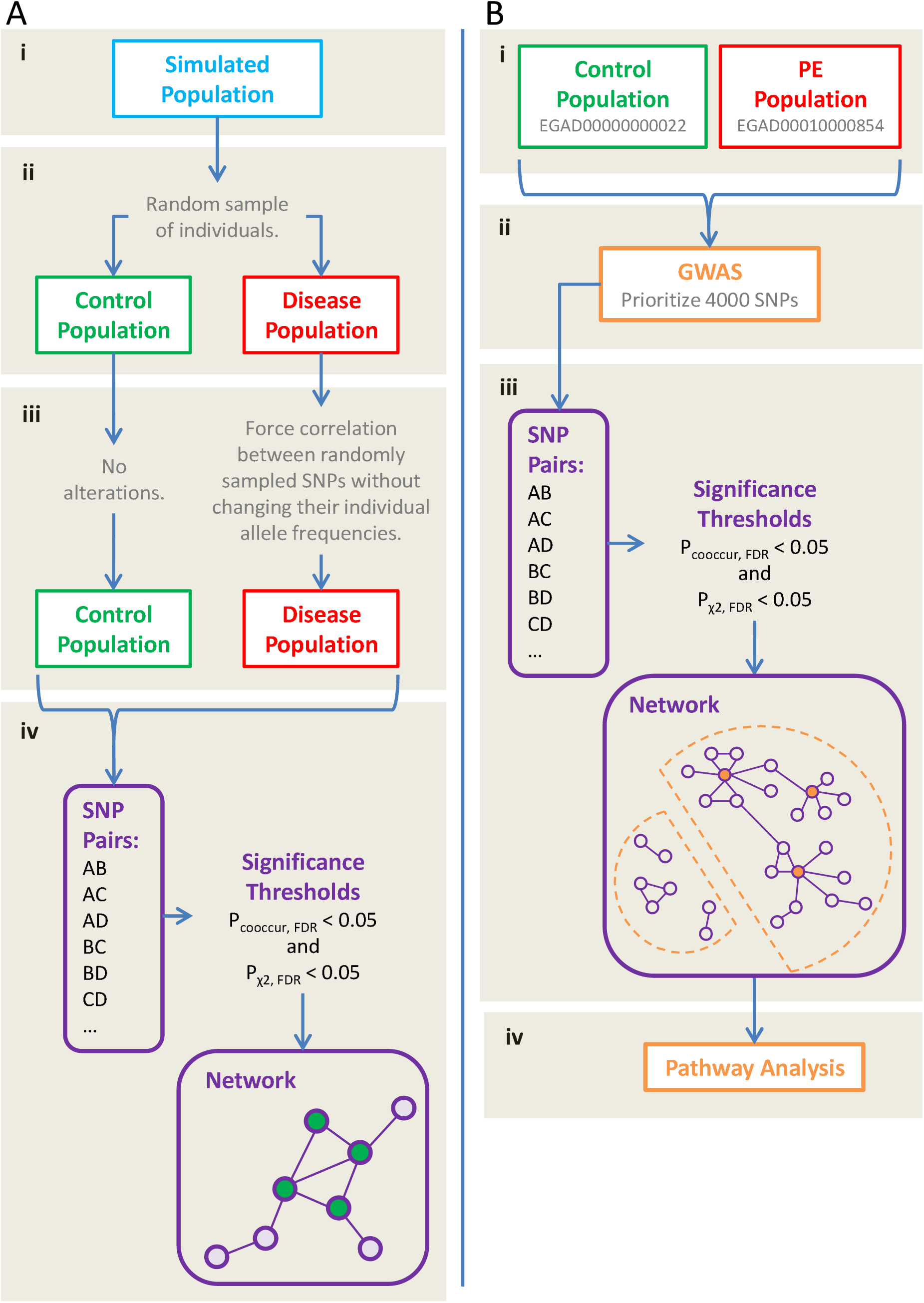
Method Overview. (A) Simulation workflow. (i) A general population of genomes is generated: 6000 individuals, each with 4000 SNPs. SNPs are coded as binary genotypes with 0s and 1s representing the absence or presence of a minor allele, respectively. At this stage, the LD parameter was varied to allow the proposed co-occurrence network to be tested on different LD models. (ii) Random sampling is used to create control and disease populations. Ten pairs of datasets are generated for a given model. (iii) In the disease population, a subset of SNPs are randomly selected and forced to correlate with one another while their frequencies are maintained. These are referred to as the spiked SNPs. Different disease models are represented by varying the number of spiked SNPs and the correlation level between them. (iv) The proposed co-occurrence method was tested. All possible pairs of SNPs were analyzed. Pairs that significantly co-occurred at significantly different rates in the two populations were placed into a co-occurrence network. (B) Workflow for real-world data. (i) Control and PE data were downloaded from EBI. (ii) Standard GWAS protocols were applied to the data for quality control. To match the input data size in simulations, 4000 SNPs with the smallest p-values from GWAS were selected for further analysis. (iii) The proposed co-occurrence method was applied as in the simulations. (iv) Pathway analysis was conducted on several SNP sets.

We observed that the models with more spiked SNPs had a higher correlation between spiked SNPs and returned a greater number of SNP pairs as statistically significant into the co-occurrence network (Wilcox test, FDR adjusted p-value ≤ 0.05). The model with five spiked SNPs at a correlation of 0.2 had, on average, 41 SNP pairs placed into the co-occurrence network. The model with 80 spiked SNPs at a correlation of 0.4 had, on average, 4003 SNP pairs placed into the network. As the number of SNP pairs passing into the network increased, the total number of SNPs increased, averaging 56 SNPs in the former model and 1033 SNPs in the latter.

Next, we graphically evaluated the models using networks (Fig. 2A-C). Each node represents a SNP, and an edge was drawn between all pairs of SNPs that passed the co-occurrence filter. For all models, the network had a structure where spiked nodes appeared to be the network’s hubs, belonging to a larger, more connected sub-network (Fig. 2A-C). Surrounding the connected sub-network are smaller disconnect sub-networks that did not contain spiked nodes/alleles.

**Figure 2.**
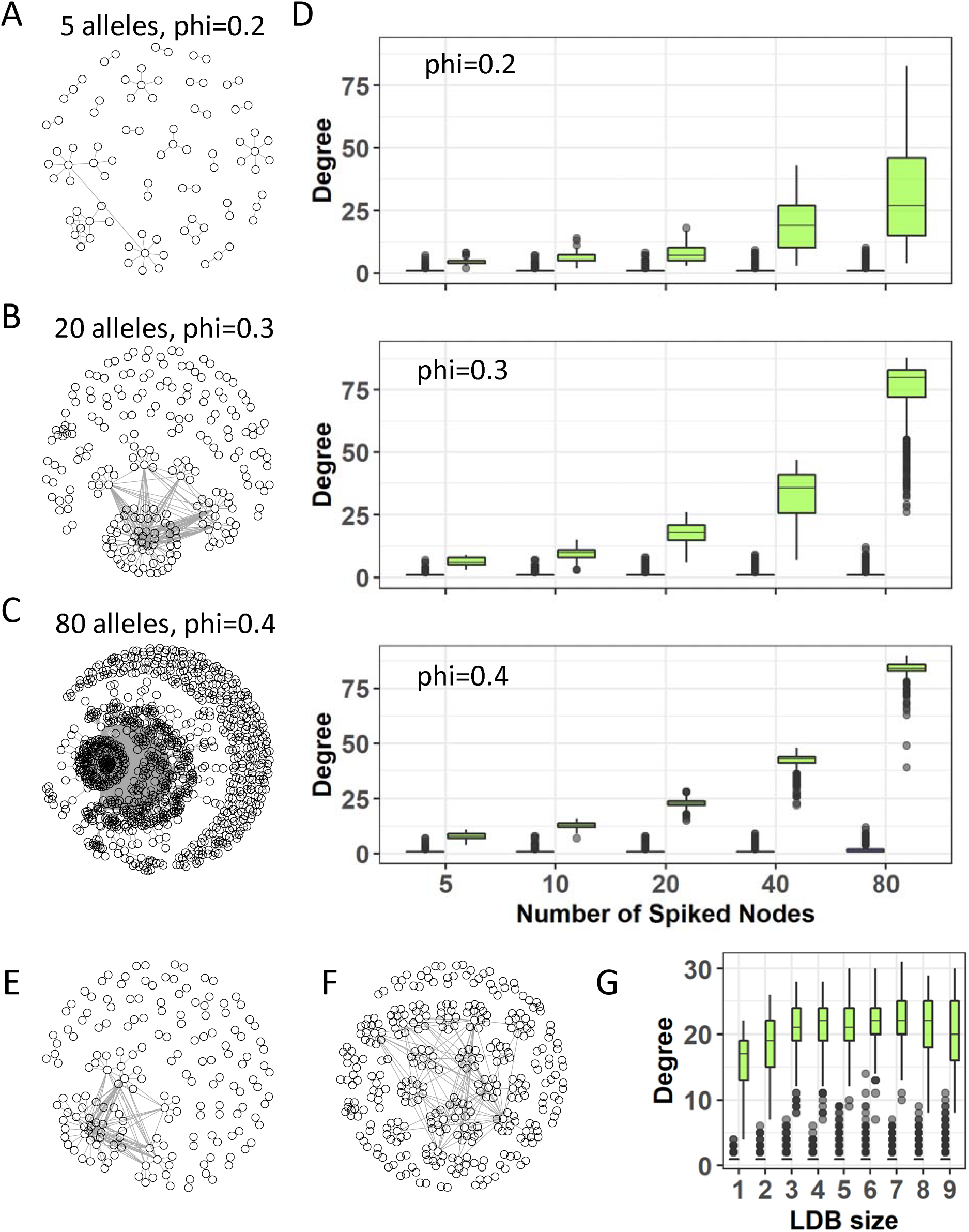
In simulations, the co-occurrence method identified SNP networks with low background. (A - C) Networks generated by disease models with 5 SNPs at a correlation of 0.2, 20 SNPs at a correlation of 0.3, and 80 SNPs at a correlation of 0.4, respectively. Nodes represent SNPs and edges link pairs that passed the co-occurrence filter. Spiked SNPs are green. Background SNPs in the connected subnetwork are orange. The connected subnetwork is the set of nodes that connect to spiked nodes, either directly as neighbours or indirectly via other nodes. Background SNPs in the disconnected subnetwork are purple. (D) relationship between the degrees of spiked nodes (green boxplots) vs. background nodes (blue boxplots) by the number of spiked SNPs from 5 to 80, representing ten trials. Each plot is a different correlation level (phi 0.2 to 0.4). At a phi=0.2 only 80 SNPs was significant(Wilcox test, FDR corrected P value so symbol< 0.05), at all other phi values all ranges of SNPs significantly different(Wilcox test, FDR corrected P value so symbol< 0.05). (E, F) Networks generated by LD models with 1 and 9 SNPs per LD block, respectively. Colours are described in A – C. (G) Degree of nodes across LD models. Each pair of boxplots represents all nodes from 10 trials of a given model. Green boxplots represent spiked nodes, and blue boxplots represent background nodes. Models are laid out left to right from one to nine SNPs per LD block.

We evaluated the graph properties of the spike nodes relative to the non-spiked (background) nodes in the network graphs. Across all models, the mode of the spiked SNPs’ degrees was significantly higher than the mode of the background SNPs’ degrees (Fig. 2D). Furthermore, the minimum degree of the spiked SNPs and the maximum degree of the background SNPs was significantly different for disease models with a correlation of 0.3 or 0.4 (Wilcox test, FDR corrected P value < 0.05). At the 0.2 correlation level, only the model with 80 SNPs had significantly different extremes for the degree of spiked and background SNPs(Wilcox test, FDR corrected P value < 0.05) (Fig. 2 D). Based on the recall properties of the spiked SNPs, we estimated a study could be powered with a minimum of 250 each case and controls for a weak correlation (phi=0.2) and 100 cases and controls for a strong correlations (phi=0.4) to exceed an 80% recall rate (Fig. S1).

### LD block size does not affect SNP co-occurrence identification

LD block size varies with genetic subpopulations, which can be problematic when making comparisons between populations. To address block size, we re-evaluated the simulated case-control models while varying the LD block size when generating the simulated genomic populations (prior to any creation of paired control and disease datasets). Network structure appeared to be relatively unaffected by variation in LD. Graphs of different LD models were highly similar, where spiked nodes were again network hubs with a higher degree than background nodes and part of a larger, more connected sub-network. (Fig. 2E-G).

### Co-occurring SNPs in preeclampsia

To evaluate the performance of our co-occurrence method on real-world data, we accessed a previously published GWAS data set on a case-control study of preeclampsia (Fig. 1B). Genotyping data was obtained from the EGA dataset repository as PLINK files (control dataset EGAD00000000022 and PE dataset EGAD00010000854). Control data was a general control population used by WTC to study other diseases and therefore contained males and additional SNPs^26^. Males were filtered out, and only SNPs genotyped in the PE population (maternal genomes) were retained. Of note is that controls were not collected to exclude PE cases, though PE has a low prevalence of about 5% worldwide^15^. Standard GWAS quality control procedures were applied with no imputation. The final dataset contained 3172 individuals (1320 control, 1852 PE) and 509,532 SNPs.

To correct and control for probes with a high degree of collinearity due to batch effects of cases and controls, we modelled the association of SNPs to patient classes was tested assuming an additive model using logistic regression with five multidimensional scaling components as covariates to account for ancestry. After correction, 93 SNPs still passed a Bonferroni significance threshold (9.8 x 10^-8^), and 350 SNPs failed due to multi-collinearity. We removed all 350 failing SNPs. This is a minimal loss relative to the over 500,000 SNPs passing QC.

### Preeclampsia genetics produce a co-occurrence network

As the co-occurrence network method was applied to data with unusually high GWAS-significant SNPs, we evaluated data sets with and without the 93 passing SNPs. We assumed that most of these Bonferroni passing SNPs represent spurious differences due to a residual batch effect of merging the two cohorts. While this was not a systematic assessment of all SNPs and does not guarantee all spurious signals were removed, it removes SNPs with the strongest univariate signals, which are the SNPs most likely to affect the co-occurrence network by adding random noise.

SNP ranking is a common approach used in polygenic risk models of GWAS data^20^. We identified the top 4000 SNPs ranked by their p-value for differential enrichment between cases and controls. Of the 4000 input SNPs, our cooccur method placed 2895 in a network of 9755 pairs that passed the co-occurrence filter. Graphing the network (Fig. S2A) showed that although the size of the network was more extensive than observed in simulations (Fig. 2A-C), the structure of the network remained consistent (Fig. S2A). The preeclampsia data network produced a larger connected sub-network and a separate set of many smaller discrete sub-networks, herein referred to as the disconnected sub-network. We observed that the disconnected sub-network was primarily composed of small (<9 nodes) self-contained modules (Fig. 3A). These were removed for clarity in the graph (Fig. 3B). As expected from our simulations, the distribution of degree in the network was bimodal (Fig. 3C and inset i), as co-occurring disease-associated SNPs should have higher degrees than other nodes in the network as indicated by our modeling (Fig. 2). Excluding the 93 GWAS-significant SNPs from the network resulted largely in removal of nodes from the disconnected subnetwork (Fig. S2B). As the disconnected network is predicted to be background we focused our investigation on the connected co-occurrence network.

**Figure 3.**
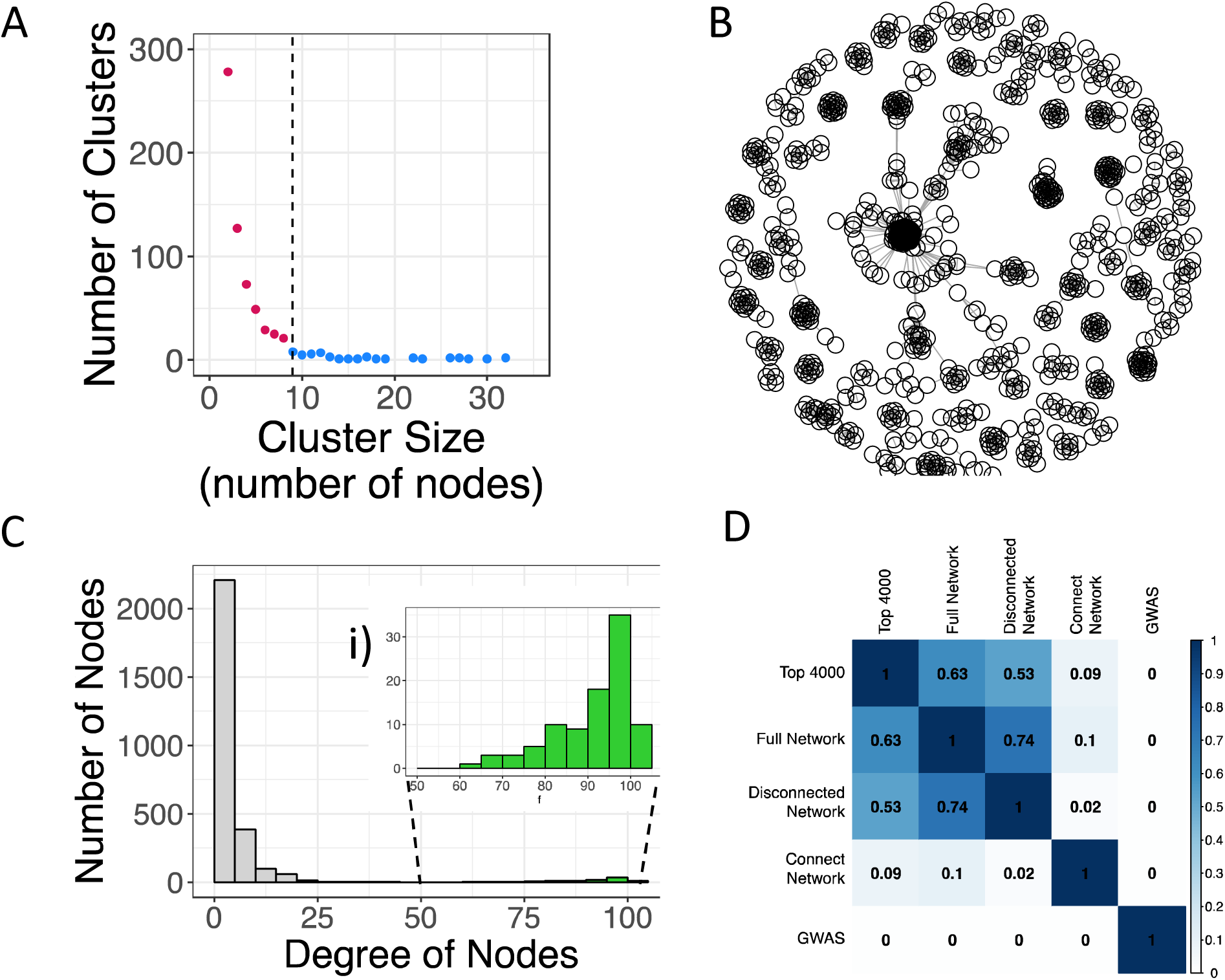
Preeclampsia genetic co-occurrence network. (A) Filtered view. Clusters of less than nine nodes in size are removed to visualize the network structure better. Nodes represent SNPs and edges link pairs that passed the co-occurrence filter. Purple represents the disconnected sub-network, and orange represents the connected sub-network. (B) Distribution of size of clusters in the full network. The dotted grey line marks clusters of size 9. The clusters greater than 9 nodes are displayed in A. (C) Distribution of degree of nodes in the full network showing a bimodal distribution (inset figure i). D) Column and row names represent the SNP sets for which pathway analysis was conducted. Numbers represent the Jaccard coefficient of overlap in significant pathways for two given SNP sets (0 means no overlap, and 1 represents complete overlap. The Top 400, Full Network and Disconnected Network all show a high level of overlap. The Connected Network is highly unique. The GWAS network is also unique but limited in total number of discovered pathways.

### Co-occurring SNP network is enriched in pathways related to PE pathophysiology

We proposed that the accumulation of minor allele SNPs within pathways leads to poor function and risk of preeclampsia. To test this, we used over-representation analysis of significantly co-occurring SNPs for the enrichment of Gene Ontology^27^. We accepted only Gene Ontology’s with a fold enrichment ≥ 2 and FDR-adjusted p-value ≤ 0.05 as significant. Critically, we evaluated multiple SNP groupings for enrichments to evaluate if the co-occurrence network contained more abundant and interpretable pathways in the context of PE pathophysiology. Specifically, we tested the full network, connected and disconnected sub-networks separately. The top 4000 and GWAS-significant SNPs were also tested for pathway enrichment. To globally assess the enrichment similarities between the different SNP sets, we evaluated the overlap of discovered pathways using a Jaccard score (Fig. 3D). The top 4000 SNPs, the entire co-occurrence network and the disconnected network were highly overlapped with a Jaccard score greater than 0.5 for all comparisons. However, the connected network was highly different, with all Jaccard scores of 0.1 or less relative to other comparisons. This was interesting as the connected network is a subset of all the top 4000 SNPs and the entire cooccurrence network.

As enriched ontologies can be highly redundant due to genes sharing multiple annotations and the hierarchical nature of the ontologies, we used the Cytoscape^28^ plugin EnrichmentMap^29^ to consolidate highly similar ontologies and create annotations based on high-frequency words in the ontology titles/descriptions^30^. The connected sub-network produced many ontological clusters that related to known aspects of PE pathophysiology (Fig. 4). Most prominent was a cluster of ontologies related to vascular endothelial growth factor (VEGF) signalling (Fig 4 and S3A). Defects in maternal vascular (spiral artery) remodelling are a hallmark of many PE cases. Although this is known, the SNPs discovered by co-occurrence may describe the genetic changes that lead to a poor maternal angiogenic response to pregnancy. Other pathways of interest involve immune functions, specifically leukocyte-mediated responses, chemokine interactions, interferon gamma signalling, antigen presentation and lymphocyte migration. We and others previously observed altered immune cell infiltration patterns in PE, including a subtype of PE that appears to be of a maternal immune system origin^12,31^. We verified these results after the removal of the 93 Bonferroni GWAS significant SNPs from the co-occurrence network. This produced a highly similar network graph enriched for the same ontologies (Fig. S3B), likely as the majority of the 93 SNPs are in the disconnected network.

**Figure 4.**
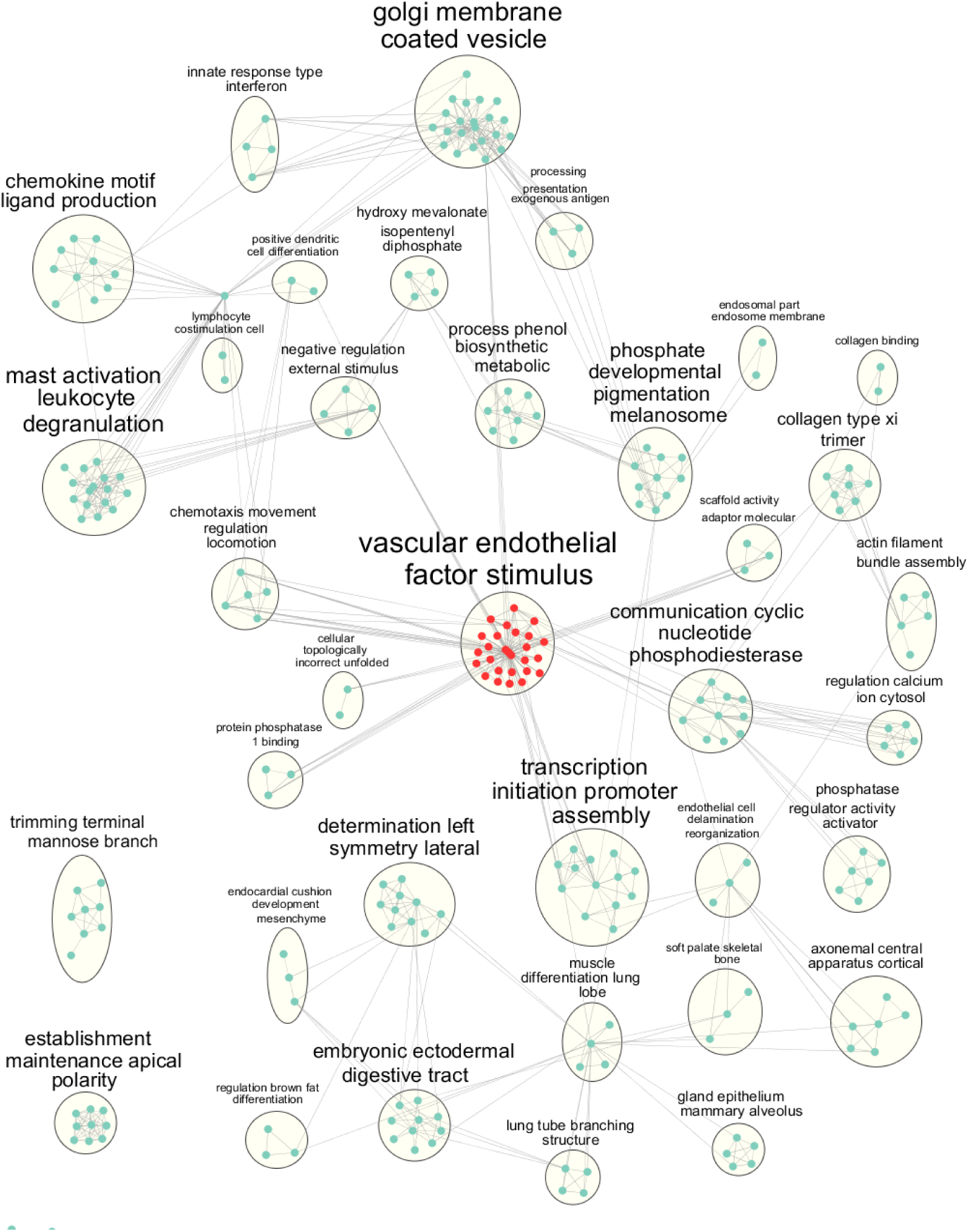
The connected sub-network is enriched in Gene Ontologies related to the PE pathophysiology. Nodes represent gene ontologies, and edges represent genetic overlap of ontologies (shared gene names). Clusters are named after the most common word patterns in the ontologies they contain. The largest cluster related to VEGF signalling is highlighted in red. All ontologies had a fold enrichment ≥ 2 and FDR-adjusted p-value ≤ 0.05.

Our modelling predicted that the disconnected network would be background co-occurrence unrelated to PE, unexpectedly we observed a high number of enriched ontologies, many with known association to preeclampsia pathophysiology (Fig. S4A). Of interest and relating to previous observations in PE patients were pathway clusters related to plasma sodium/potassium transport, phagosome pH, atrial action, urea and creatine homeostasis, and steroid biosynthesis, including glucosylceramides and thyroid stimulation.

However, a standard GWAS did not produce many enriched ontologies (data not shown). The top 4000 SNPs and the full network did not produce as many ontologies as the disconnect or the connect networks, despite a larger number of genes (Fig. S4B).

## Discussion

Little progress has been made on the treatment and predictive diagnosis of preeclampsia^2^. Likely this results from the high heterogeneity between patients due to subtypes of the disease observed by us and others^12,32–35^. We modelled PE genetics as sets of interacting SNPs/alleles that frequently co-occur in PE to improve predictive testing and spur new avenues for PE therapy development.

Our modelling of the co-occurrence method indicated that bona fide co-occurring SNPs would form networks with high degrees and interconnectivity. Spurious co-occurrence tended not to be connected to the high-degree network and is composed of low-degree nodes. This finding fits our proposal that these multigenic causes of PE are due to the accumulation of minor alleles within pathways. Encouragingly we observed an overall similar graph topology using real-world PE data, with a highly connected central SNP network and smaller discrete networks disconnected from the central network. The agreement between the model data and the real-world data suggests that the method is working to identify SNPs associated with PE.

Significantly the connected network was highly enriched in biological processes with many related to the pathophysiology of PE. These included VEGF signalling. Poor VEGF signalling and angiogenesis are frequently observed as a feature of pathology, circulating biomolecules and genetic associations^13,15,19,36,37^. Other enriched ontologies of interest were related to immune cells and the presentation and trafficking of antigens. Paternal antigens are thought to play a role in PE risk for first pregnancies^38^. Our previous analysis of PE cohorts identified a maternal immune subtype with molecular and histopathology signs of graft versus host disease^12,13,31^. Generally, subsequent pregnancies with the same partner are at a lower risk due to maternal immune priming or adaptive tolerance to the paternal epitopes/antigens^22^.

Unexpectantly, the outer disconnected network was enriched in gene ontology terms related to possible roles in PE. If disconnected network were random noise, we would expect few enriched ontologies. Ontology enrichments of the disconnected network that may be related to PE pathophysiology were sodium-potassium plasma regulators, atrial action^39^, urea creatine homeostasis^40^, steroid biosynthesis and thyroid stimulation^41^. A possibility is that the disconnected network may contain genetic co-occurrence networks for associated comorbidities. Pre-existing conditions such as chronic hypertension and obesity are considered risk factors for PE but can occur independently of PE^42^. Potentially some forms of preeclampsia are the accumulation of a series of comorbidities that reduce the ability of the mother’s physiology to adapt to pregnancy. The genetic presence of a comorbidity may synergize with PE causal SNPs. A more rigorous case-control design that balanced obesity and other patient demographics could test this hypothesis.

An additional explanation for the enrichment of ontologies to the disconnected network is that the network is incomplete. As we used only 4000 ranked SNPs, using more SNPs might produce a larger co-occurrence network that links members of the disconnected network to the connected network. The poorer performance of ontology enrichment analysis using either the top 4000 SNPs or the full network is likely due to false positives let in by the permissive p-value in the absence of a second test (such as co-occurrence).

We encountered a few hundred probes with spurious correlation to patient classes likely due to residual batch effects, which further resulted in 93 SNPs passing a Bonferroni correction using standard GWAS method. However, the potentially spuriously passing SNPs that reached the cooccurrence network generally fell within the disconnected network. Our simulation models indicate that the disconnected network is background noise. The identification of the spuriously passing SNPs in the disconnected network indicates that our method may be robust to noisy data, potentially due to the application of two significance tests and the demand of cooccurrence embedded in a higher order structure of the connected network.

Importantly, the results of our study remaining to be reproduced in other cohorts of PE cases. Similar to typical GWAS findings, we cannot conclude that the observed co-occurrence SNPs are causal to the pathology of PE but may be linked to the causal SNPs. A possible approach is the imputation of genotypes to resolve the potential causal alleles. Our method may be applicable to sequencing-based variant discovery, such as exome resequencing or genome sequencing, but we have not validated this approach. Currently, we are working to improve this method to enable a more significant number of alleles to be tested for co-occurrence and to identify genetic subtypes of disease.

The results of our analysis may form the basis of genetic tests before or early in pregnancy. In the future, a combinatorial genetic test could classify a patient into PE subtypes and identify specific molecular and biochemical susceptibilities. Knowledge of the disrupted pathways is a start to personalized medicine approach to treating PE. Broadly this method could be applied to any disease or trait to identify co-occurrence genetic networks as the method uses standard SNP chip data sets. Other conditions known to be polygenic, including cardiovascular disease or neuropsychiatric disorders, may benefit from applying our co-occurrence method to existing data sets. To facilitate the application of the co-occurrence method to other data sets, we made the code available through a GitHub repository with supporting documents.

## Methods

### Co-occurrence filter and network

For case-control genomic data, pairs of SNPs passing two co-occurrence filters created networks. The first filter selected SNP pairs that co-occurred more or less frequently than expected in the control or case population. Co-occurring refers to at least one minor allele at both SNP loci. Co-occurrence was calculated by a customized version of the co-occur function from the R package co-occur, which uses a hypergeometric test^43^. SNP pairs with FDR-adjusted p-values ≤ 0.05 passed the filter. We parallelized the function to run more efficiently but limited all analyses to a maximum of 4000 input SNPs. The second filter selected for SNP pairs that co-occurred at significantly different rates in control vs case populations. A chi-squared test was used, and SNP pairs with FDR-adjusted p-values ≤ 0.05 passed the filter. Networks, with nodes representing SNPs and edges representing co-occurrences, were created with the graph package in R^44^.

### Simulations for method validation

For validation, the co-occurrence method was applied to artificial case-control populations. A population of artificial genomes was generated with the R function to generate SNPs from the package minutest (version 1.7)^45^. Genotypes were re-coded to 0s or 1s to represent minor alleles’ absence or presence. A population of 6000 subjects was generated. To generate a single trial of a case-control dataset, two sets of 1000 genomes were sampled from the population to form the case and control cohorts without replacement. Before altering the case-cohort, the co-occurrence method was applied to assess its results when no difference was expected.

In the case-cohort, the correlation was forced between a set of randomly sampled SNPs (herein called spiked SNPs). Individual frequencies of the SNPs were maintained to ensure no difference in individual SNP frequency between case and control. This was accomplished with the function rmvbin from the R package bindata^46^. When provided with a matrix containing the desired correlation level between sampled SNPs and the minor allele frequencies of the sampled SNPs, the function generates genotypes (0s and 1s) that satisfy both the desired correlation and minor allele frequencies. These genotypes replaced the original genotypes of the sampled SNPs in the disease population. In total, ten cohorts were generated.

Fifteen disease models were generated by combinations of the number of spiked SNPs (5, 10, 20, 40, or 80), and the correlation level (0.2, 0.3, or 0.4) was varied. For the disease models, the background genomic structure of the population was kept constant (2 SNPs per LD block, 8 LD blocks per gene, and 250 genes). For the disease models, ten pairs of case-control were drawn from the population (before spiking SNPs). Each of the 15 disease models that varied SNP number and correlation value was built from the same ten cohorts.

Additionally, nine LD models were generated, where the number of SNPs per LD block varied from 1 to 9. The number of genes was set to keep the size of the simulated genome close to but under 4000 SNPs. The disease model used for LD models was kept constant (20 SNPs at a correlation of 0.3). As the population parameters were altered during the simulation, a new set of 10 case controls for each LD model were used for each LD block size.

Individual SNPs were tested for disease association using the Cochrane-Armitage trend test (p ≤ 5 x 10^-8^) to demonstrate that the generated case-control datasets did not yield significant univariate results. The co-occurrence method described above was then applied to the datasets. The resultant networks were then assessed. Spiked nodes were coloured differently in the network to determine if they were visually distinguishable from background nodes. Their degree (number of neighbouring nodes in the network) was assessed. A Wilcox test was used to determine if the mode of the ten cohorts spiked SNPs’ degrees significantly differed from the mode of background SNPs’ degrees (FDR adjusted p-value ≤ 0.05) within a given disease or LD model. Likewise, a Wilcox test was used to determine if the minimum of spiked SNPs’ degrees significantly differed from the maximum of background SNPs’ degrees (FDR adjusted p-value ≤ 0.05).

### Preeclampsia data: QC, GWAS, and co-occurrence

Genotyping data was obtained from the European Genome-Phenome Archive’s dataset repository. The Wellcome Trust Case Control Consortium (WTC) originally collected the data. The PE dataset (EGAD00010000854) containing maternal genomic data and the control dataset (EGAD00000000022) were collected separately though both cohorts were from the United Kingdom. The controls served as common controls for the WTC to study several diseases. They thus included males and were not described as excluding PE. However, PE has a low prevalence of around 5%^8,15^.

A standard GWAS protocol, using PLINK^47^, KING^48^, and R, was applied to the data for quality control and to prioritize a set of 4000 SNPs to input into the co-occurrence method. Control and disease data were merged, and only SNPs in both datasets were kept. Only females were kept. No imputation was done as the co-occurrence method was applied to only 4000 SNPs. The following quality control measures were performed: individuals with discordant sex, genotype missing rates ≥ 0.025, heterozygosity ≥ 2 standard deviations away from the mean, and non-northern European ancestry (inferred using KING) were removed. One individual from every related pair (IBS > 0.185) was removed. SNPs with genotype missing rates ≥ 0.05, with genotype missing rates that differed between control and PE (FDR adjusted p-value from chi-squared test ≤ 0.05), that deviated from Hardy-Weinberg equilibrium (p ≤ 1.0 x 10^-6^), and whose minor allele frequency was ≤ 0.05 were removed. Association was tested assuming an additive model using logistic regression with five multidimensional scaling components. Bonferroni significance level was used (0.05 divided by the number of SNPs).

Only 4000 SNPs were input into the co-occurrence method to remain consistent and allow comparison to simulations. The 4000 SNPs with the smallest p-values from the GWAS were used. Genotypes of these SNPs were coded as 0s and 1s, and the co-occurrence method described earlier was then applied.

### Pathway enrichment of genotypes in co-occurrence network

Pathway enrichment was conducted using GREAT^49^. This web-based tool assigns SNPs (including intergenic ones) to genes and then tests for their over-representation in pathways. The set of all SNPs passing quality control was used as the genomic background for these tests. The association rules used to assign SNPs to genes were left as their default: “basal plus extension” with proximal set to 5.0 kb upstream and 1 kb downstream, distal set to 1000 kb, and curated regulatory domains included. Gene Ontology pathways with fold enrichment ≥ 2 and FDR-adjusted p-value ≤ 0.05 were considered significant.

Cytoscape 28(version 3.7.2) was used to interpret the many pathways returned^28^ to create a less redundant network where pathways (nodes) with high genetic overlap are linked. Within Cytoscape, the Enrichment Map^29^ and Auto-Annotate^30^ apps were used to create the network and summarize its clusters.

## Data Availability

Genotyping data was obtained from the European Genome-Phenome Archive's dataset repository. The Wellcome Trust Case Control Consortium (WTC) originally collected the data. The PE dataset (EGAD00010000854) containing maternal genomic data and the control dataset (EGAD00000000022) were collected separately, though both cohorts were from the United Kingdom.

## Acknowledgments

High performance computing infrastructure and technical support was provided by Compute Canada and SciNet.

The authors that they have no competing financial interests and that funders had no input into research or review of publication of results.

---

**Supplemental Figure 1.**
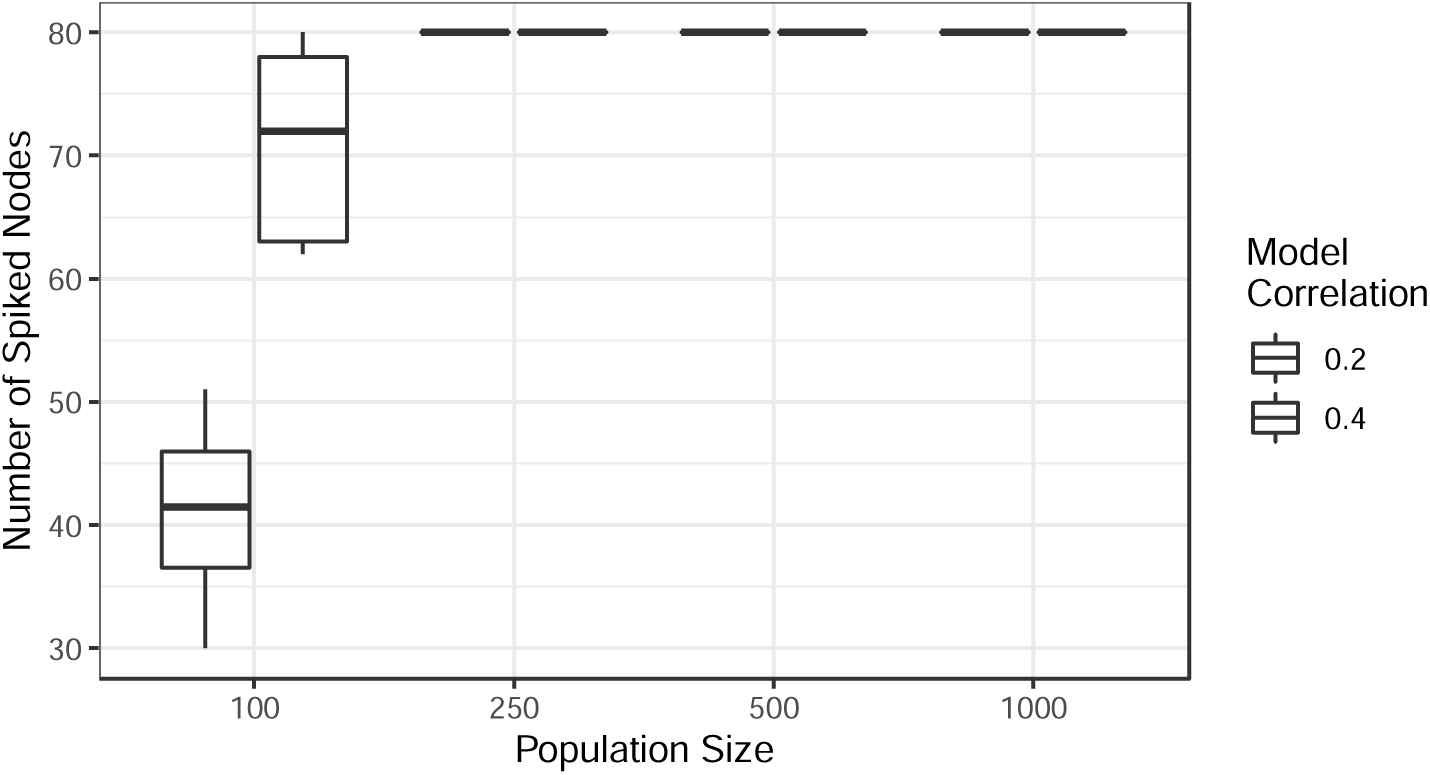
Power analysis simulation. Power was empirically estimated by the ability to recall 80 spiked nodes in a simulation study. Ten simulations were performed for populations of 100-1000 at phi 0.2 (weak effect) and 0.4 (strong effect).

**Supplemental Figure 2.**
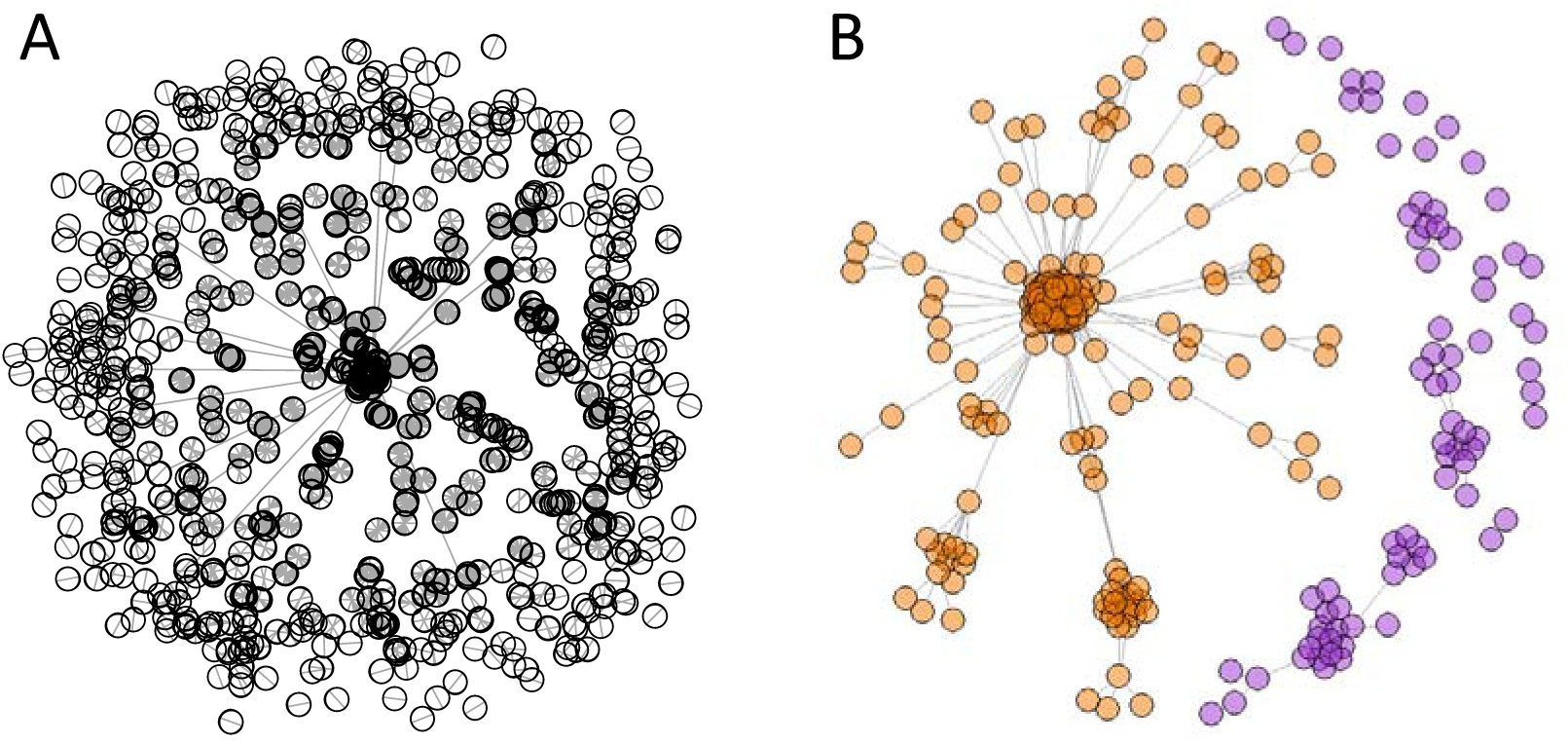
Other visualizations of the PE co-occurrence network. (A) Unfiltered network. Nodes represent SNPs and edges link pairs that passed the co-occurrence filter. Purple represents the disconnected sub-network, and orange represents the connected sub-network. All nodes are shown, including small subnetworks less than none nodes; shown are 2859 nodes connected by 9755 edges. Nodes overlap due to the large number of nodes, making the network structure challenging to see. (B) Connected sub-network after removal of GWAS-significant SNPs. Nodes that lost their connection to the main connected sub-network have been re-coloured purple.

**Supplemental Figure 3.**
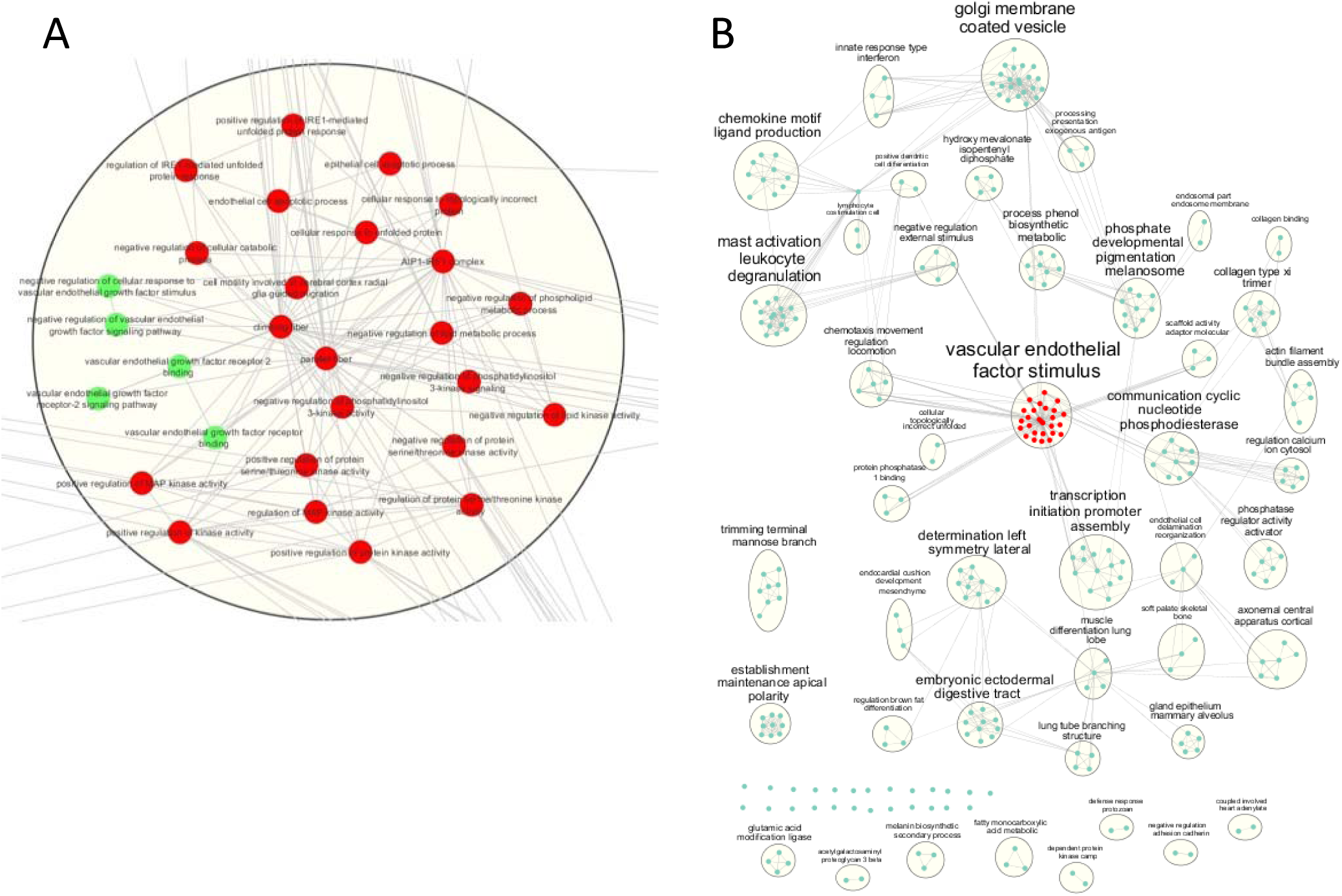
VEGF signaling-centered network. A) detailed view of the pathways with significant gene overlap and annotated to VEGF signalling and related pathways. B) Enriched Gene ontologies after removal of the GWAS passing SNP/genes. A minimal change in enrichment is observed.

**Supplemental Figure 4.**
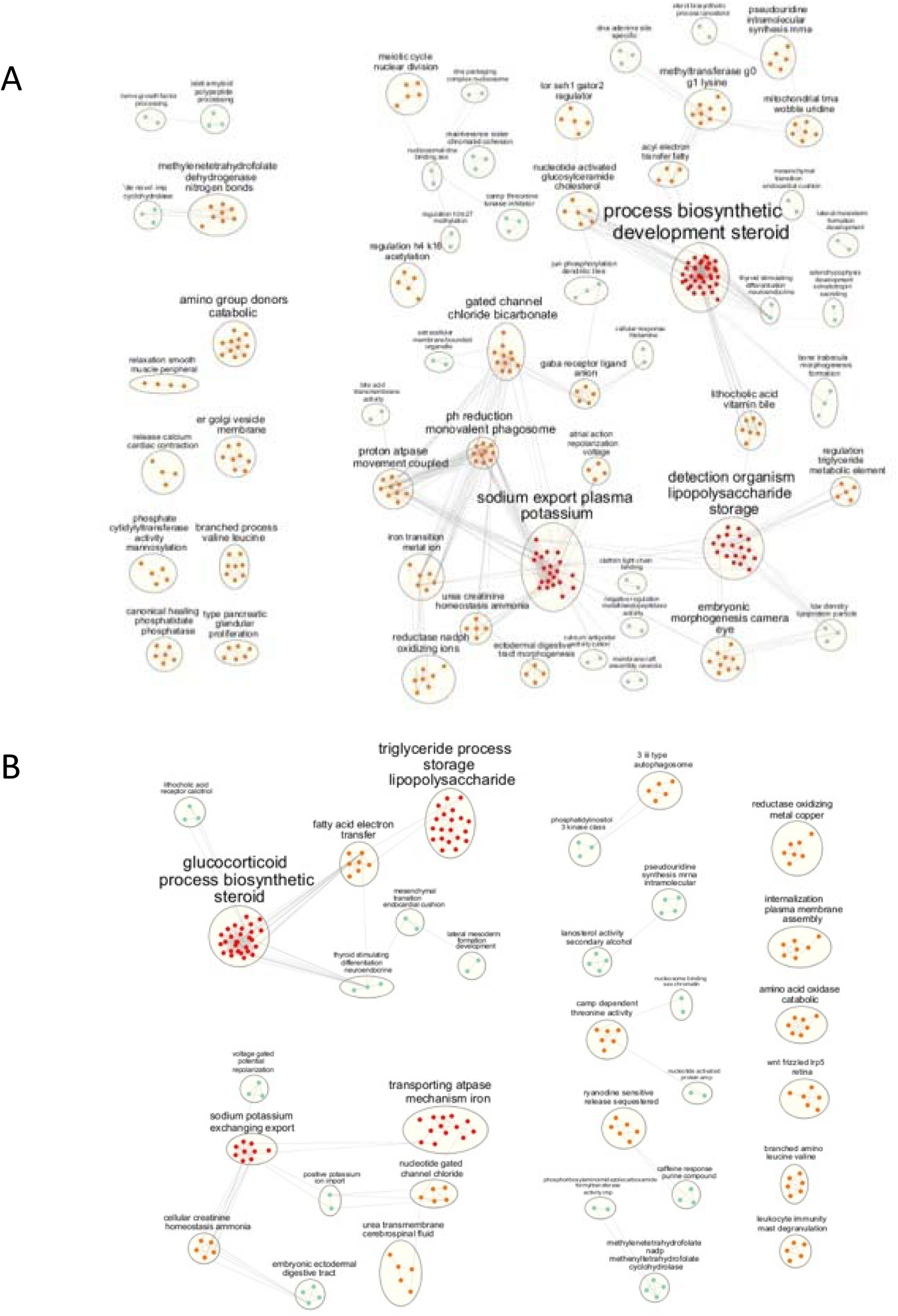
The disconnected network is enriched in ontologies related to the pathophysiology of preeclampsia. A) The enrichment map of the ontologies is significantly associated with the SNPs of the disconnected network. Many are known to be related to the pathophysiology of preeclampsia. B) The enrichment map of the top 4000 SNPs used to build the co-occurrence networks.

