## supplemental figures for "Genetic co-occurrence networks identify polymorphisms within ontologies highly associated with preeclampsia"

### Fig S1

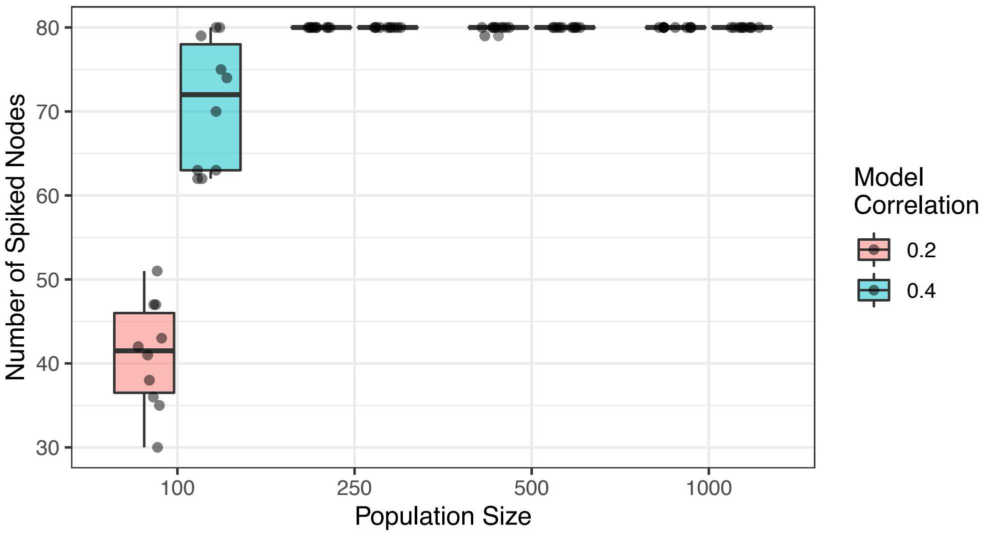


**Supplemental Figure 1. Power analysis simulation.** Power was empirically estimated by the ability to recall 80 spiked nodes in a simulation study. Ten simulations were performed for populations of 100-1000 at phi 0.2 (weak effect) and 0.4 (strong effect).

### Fig S2

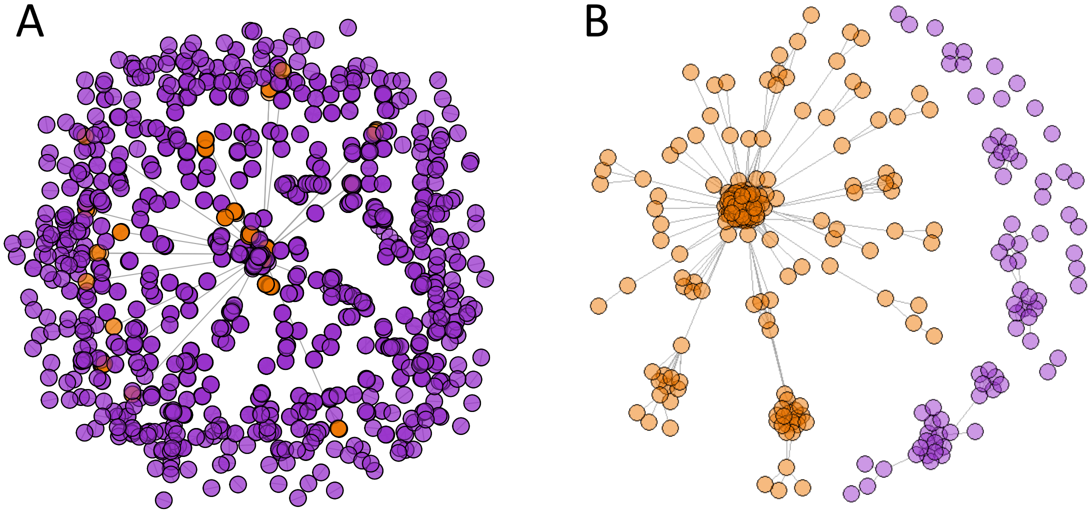


**Supplemental Figure 2. Other visualizations of the PE co-occurrence network**. (A) Unfiltered network. Nodes represent SNPs and edges link pairs that passed the co-occurrence filter. Purple represents the disconnected sub-network, and orange represents the connected sub-network. All nodes are shown, including small subnetworks less than none nodes; shown are 2859 nodes connected by 9755 edges. Nodes overlap due to the large number of nodes, making the network structure challenging to see. (B) Connected sub-network after removal of GWAS-significant SNPs. Nodes that lost their connection to the main connected sub-network have been re-coloured purple.

Fig S3

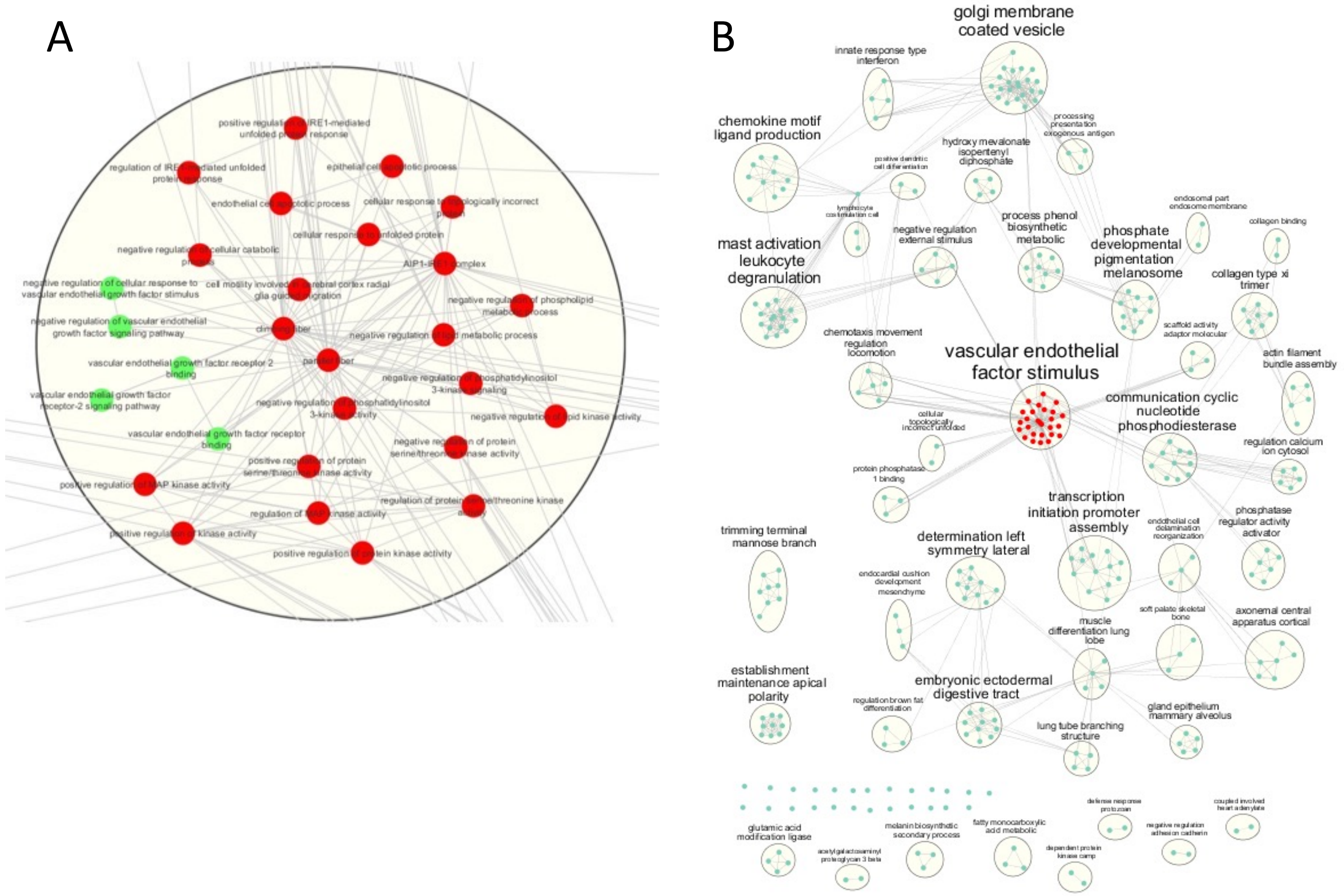


**Supplemental Figure 3. VEGF signaling-centered network.** A) detailed view of the pathways with significant gene overlap and annotated to VEGF signalling and related pathways. B) Enriched Gene ontologies after removal of the GWAS passing SNP/genes. A minimal change in enrichment is observed.

### Fig S4

A

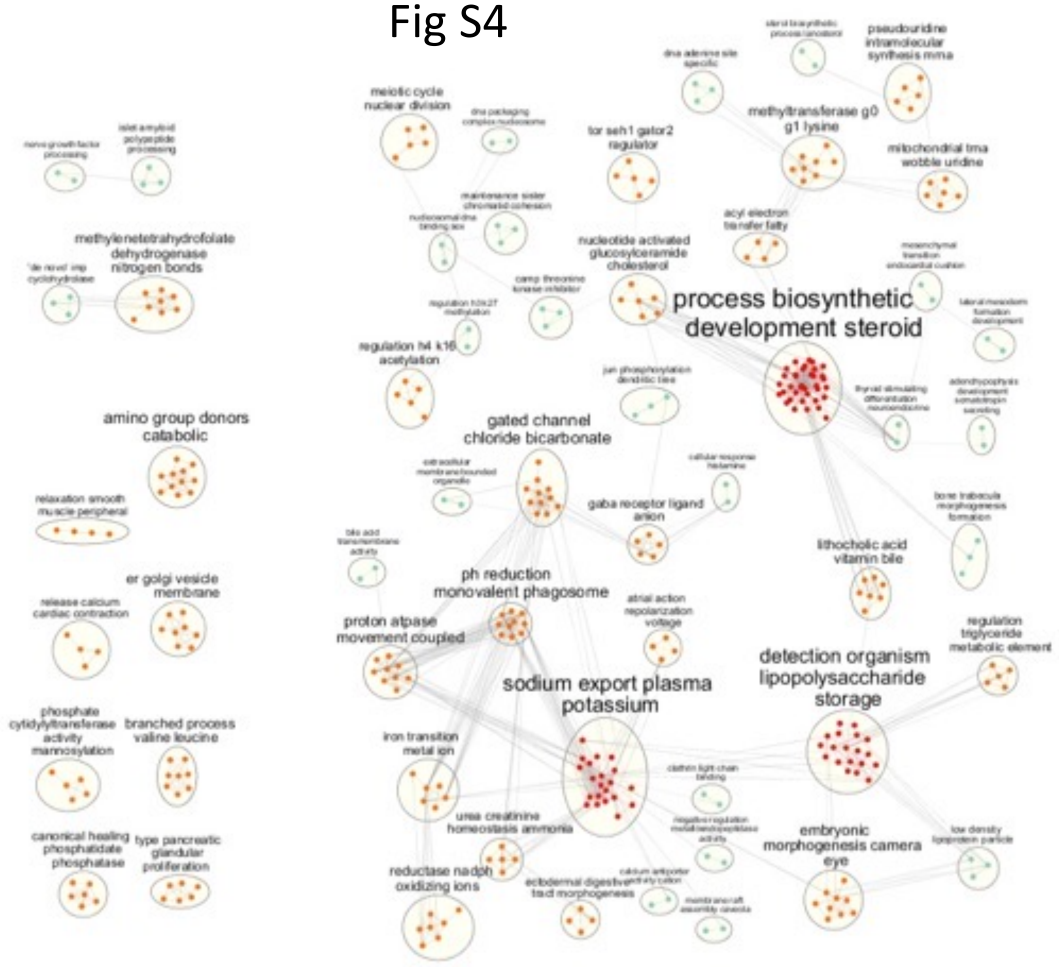


B

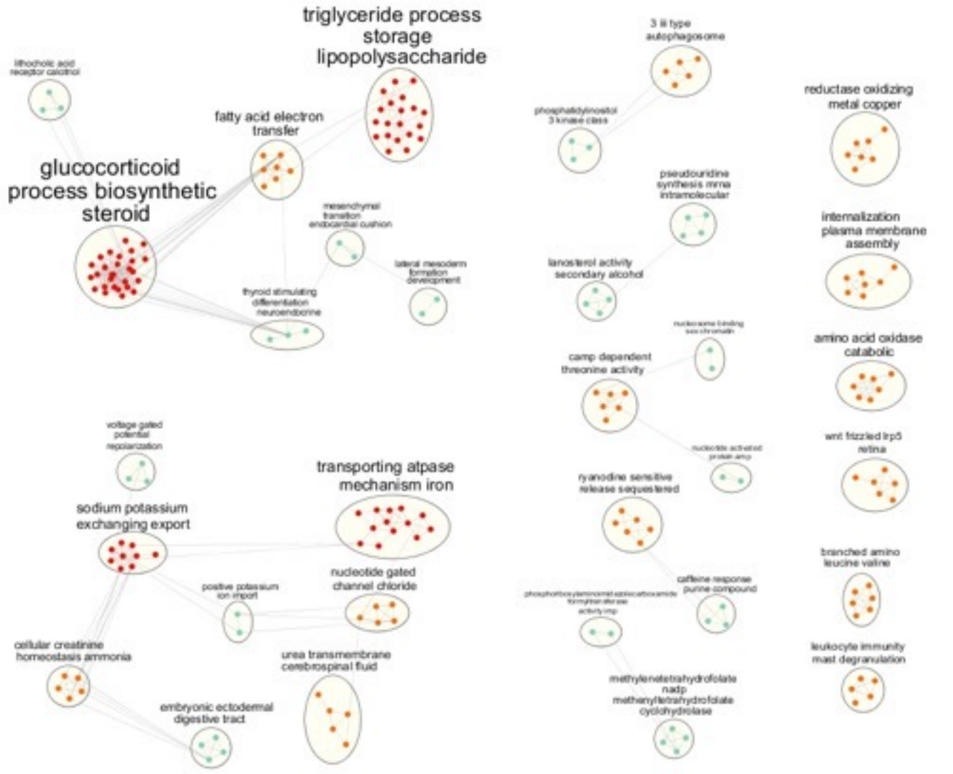


**Supplemental Figure 4. The disconnected network is enriched in ontologies related to the pathophysiology of preeclampsia.** A) The enrichment map of the ontologies is significantly associated with the SNPs of the disconnected network. Many are known to be related to the pathophysiology of preeclampsia. B) The enrichment map of the top 4000 SNPs used to build the co-occurrence networks.
